# Catechol-O-methyltransferase (COMT) Val158Met Polymorphism and susceptibility to alcohol dependence

**DOI:** 10.1101/2020.04.26.20081273

**Authors:** Amrita Chaudhary, Pradeep Kumar, Vandana Rai

**Author notes:** **Corresponding author**: Dr Vandana Rai, Department of Biotechnology, VBS Purvanchal University, Jaunpur.

## Abstract

Catechol-O-methyl transferase (COMT) enzyme catalyzes the metabolism of dopamine and other catechols in the brain. Several articles investigated catechol-O-methyltransferase (COMT) Val158Met polymorphism as risk factor for alcohol dependence (AD) but the results were inconclusive. The aim of present meta-analysis was to evaluate the association of Val158Met (COMT) polymorphism with AD. Authors performed keyword search of the four electronic databases-Pubmed, Google Scholar, Springer Link and Science Direct databases up to December 31,2019. Total eighteen studies that investigated the association of Val158Met polymorphism with AD were retrieved. The pooled results from the meta-analysis (2,278 AD cases and 3717 healthy controls) did not show association with AD using all five genetic models (allele contrast model: OR = 1.02, 95% CI= 0.90-1.14, p= 0.03; homozygote model: OR = 1.06, 95% CI= 0.81-1.38, p= 0.69; dominant model: OR = 0.99, 95% CI= 0.85-1.14, p= 0.87; co-dominant model: OR = 0.97, 95% CI= 0.86-1.11, p= 0.71; recessive model: OR = 1.05;95% CI= 0.85-1.29, p=0.61). Results of subgroup analysis showed that Val158Met is not risk for AD in Asian and Caucasian population. In conclusion, COMT Val158Met is not a risk factor for alcohol dependence.

## Introduction

Alcohol dependence (AD) is a common, chronic, relapsing neuropsychiatric disorder and isone of the leading causes of the global burden of disease (Koob 2003; Volkow *et al*. 2009; WHO, 2009). It has been demonstrated that among those who drink alcohol, only a minority (~15%) eventually become dependent on it (Hasin et al., 2007). Heritability estimates are as high as 40-60%, with involvement of multiple genes (Stacey *et al*., 2009; Bienvenu *et al*., 2010). Chronic alcoholics have suffered with different psychiatric comorbidities, such as antisocial personality disorder, anxiety, depression, and attention deficit hyperactivity disorder (Ducci et al. 2007). Dopamine is a main neurotransmitter in the development of addiction behaviour (Hyman et al., 2006). Catechol-O-methyl transferase (COMT) enzyme catalyzes the metabolism of dopamine and other catechols in the brain (Guldberg and Marsden, 1975; Mannisto et al., 1999). In the striatum and nucleus accumbens, dopamine is removed from the synaptic cleft by reuptake through dopamine transporter (DAT) and subsequent oxidation by monoamine oxidase (Eisenhofer et al., 2004). In prefrontal cortex (PF), COMT mainly controls the dopaminergic transmission, because DAT density is low in PF (Moron et al., 2002; Yavich et al., 2007). So, the gene repeatedly implicated in PFC dopamine function is *COMT*. COMT enzyme metabolizes dopamine, adrenalin and noradrenalin, and is the main factor controlling dopamine levels in the PFC.

COMT gene is present on chromosome 22q11.2, contains six exons, and expresses at high levels in many tissues including the brain, liver, kidney, and breast etc. Several polymorphisms are reported in the COMT gene, but the most studied and cinicay significant is a single base pair G to A substitution at position 472 in exon 4 (G472A), results in substitution of valine by methionine (Val158Met) in COMT enzyme (Lotta et al., 1995; Lachman et al., 1996). This substitution reduced the COMT enzyme activity. The two alleles are referred to as Val(G) and Met(A). Val allele encodes the thermostable high activity COMT enzyme and Met allele encodes the thermolabile low activity COMT enzyme (Spielman and Weinshilboum, 1981; Lotta et al., 1995; Nobile et al., 2010). Both the alleles are co-dominant, i.e. heterozygous individuals (Val/Met) have an intermediate level of COMT activity (Lotta et al., 1995). COMT gene Val158Met is a clinically functional polymorphism, and reported as risk factor for several disorders/diseases like schizophrenia (Chen et al 1997), Autism (Gadow et al., 2009), bipolar disorder (Lachman et al 1996), unipolar disorder (Ohara et al 1998), obsessive-compulsive disorder (Karayiorgou et al 1997), attention-deficit hyperactivity disorder (Retz et al., 2008), and Posttraumatic stress disorder (Valente et al., 2011).

The first study that reported an association between the COMT Val108/158Met polymorphism and substance abuse was by Vandenbergh et al. (1997). They suggested that the high COMT activity allele (Val) was linked to polysubstance abuse in Caucasians. After that several articles reported the influence of the human COMT polymorphism on alcohol/substance addiction but results remains unclear. Hence, in present study authors carried out a meta-analysis to evaluate association between COMT Val108/158Met polymorphism and alcohol dependence/addiction.

## Method

Meta-analysis was carried out according to meta-analysis of observational studies in epidemiology (MOOSE) guidelines (Stroup et al., 2000).

### Retrieval strategy and selection criteria

Articles were retrieved through Pubmed, Springer Link, Google Scholar and Science Direct databases up to December 31, 2019, using following key words: ‘Catecho-O-methyltransferase’ or ‘COMT’ or ‘Val158Met’ and ‘ Alcohol dependence’ or ‘AD’ or ‘Addiction’.

### Inclusion and exclusion criteria

Studies were included if they met the following criteria: (1) investigate the association between COMT Val158Met polymorphism and alcohol dependence risk, (2) studies with complete information of COMT Val158Met genotype/ allele numbers in alcohol dependence cases and controls and (3) sufficient information for calculating the odds ratio (OR) with 95% confidence interval (CI). Major reasons for studies exclusion were as follows: (1) no alcohol dependence cases analyzed, (2) the Val158Met polymorphism details information missing, and (3) duplicate article.

### Data extraction

The following information was extracted for each eligible study using standard protocols: (i)first author’s name, (ii)country name, (iv) year of publication, (v) journal name, (vi) number of cases and controls, and (vii) number of genotypes in cases and controls.

### Statistical Analysis

All analysis were done according to the method given in Rai et al (2014). Pooled ORs with 95 % confidence intervals (CIs) were calculated using different genetic models. Heterogeneity was investigated using Q test and quantified by I^2^ statistic (Higgins and Thompson,2002). Both fixed effect and random effect models were used to calculate ORs with their 95 % CIs (Mantel and Haenszel, 1959 DerSimonian and Laird, 1986), but model adopted on the basis of heterogeneity. in this meta-analysis. Chi-squared analysis was used to determine whether the genotype distribution of control group was in Hardy–Weinberg equilibrium or not. Subgroup analysis was conducted by ethnicity. Publication bias was investigated by using the funnel plot of precision (1/standard error) by log odds ratio and also by Egger’s regression intercept test (Egger et al., 1997). P value <0.05 was considered statistically significant. All statistical analyses were done using MIX version 1.7 (Bax et al., 2006) and MetaAnalyst (Wallace et al., 2013) program.

## Results

Initial search of four databases, 45 articles were retrieved, but 16 articles did not meet the inclusion criteria after reviewing abstract. The excluded articles include results of drug treatments of AD, book chapters, reviews, and articles investigated other genes. Out of remaining 29 articles, we also excluded thirteen articles (duplicate articles, meta-analysis and only case studied). After applying inclusion/exclusion criteria, total 16 articles (Figure 1) were suitable for the present meta-analysis (Ishiguro et al., 1999; Tiihonen et al., 1999; Hallikainen et al., 2000; Nakamura et al., 2001; Kohnke et al., 2003; Kweon et al., 2005; Enoch et al., 2006; Sery et al., 2006; Voicey et al., 2011; Wang et al., 2011; Altintoprak et al., 2012; Franke et al., 2012; Schellekens et al., 2012; Soyka et al., 2013; Malhotra et al., 2016; Carra et al., 2017) (Table 1). One author (Hallikainen et al., 2000) has studied cases from two population (Turku and Kuopio) and also one case group of type I alcoholics, so we included a these three case groups separate in the meta-analysis. Hence, in present meta-analysis, total number of included studies are eighteen. Figure 1 showed the flow diagram of study selection.

**Table 1.** Details of eighteen included studies

| **Studies** | **Country** | **Number of Cases** | **Number of Controls** | **Case Val Allele** | **Case Met Allele** | **Control Val Allele** | **Control Met allele** |
| --- | --- | --- | --- | --- | --- | --- | --- |
| Ishiguro et al., 1999 | Japan | 175 | 346 | 244 | 106 | 480 | 212 |
| Tiihonen et al., 1999 | Finland | 123 | 267 | 97 | 149 | 264 | 270 |
| Hallikainen et al., 2000, Type 2alcoholism | Finland | 62 | 267 | 64 | 60 | 264 | 270 |
| Hallikainen et al., 2000, Turku | Finland | 67 | 267 | 55 | 79 | 264 | 270 |
| Hallikainen et al., 2000, Kuopio | Finland | 56 | 267 | 42 | 70 | 264 | 270 |
| Nakamura et al., 2001 | Japan | 91 | 112 | 132 | 50 | 153 | 71 |
| Kohnke et al., 2003 | Germany | 100 | 100 | 96 | 104 | 93 | 107 |
| Kweon et al., 2005 | Korea | 97 | 94 | 144 | 50 | 132 | 56 |
| Enoch et al., 2006 | USA | 82 | 86 | 118 | 46 | 110 | 62 |
| Sery et al., 2006 | Czech Republic | 399 | 400 | 413 | 385 | 389 | 411 |
| Voisey et al., 2011 | Australia | 226 | 240 | 319 | 133 | 304 | 176 |
| Wang et al., 2011 | China | 107 | 214 | 162 | 52 | 308 | 120 |
| Altintoprak et al., 2012 | Turkey | 110 | 330 | 139 | 81 | 399 | 261 |
| Franke et al., 2012 | Netherland | 110 | 99 | 97 | 123 | 97 | 101 |
| Schellekens et al., 2012 | Netherland | 110 | 98 | 99 | 121 | 96 | 100 |
| Soyka et al., 2013 | Germany | 125 | 296 | 122 | 128 | 283 | 309 |
| Malhotra et al.,2016 | India | 210 | 200 | 214 | 206 | 237 | 163 |
| Carra et al.,2017 | Italy | 28 | 34 | 31 | 25 | 42 | 26 |

**Figure 1.**
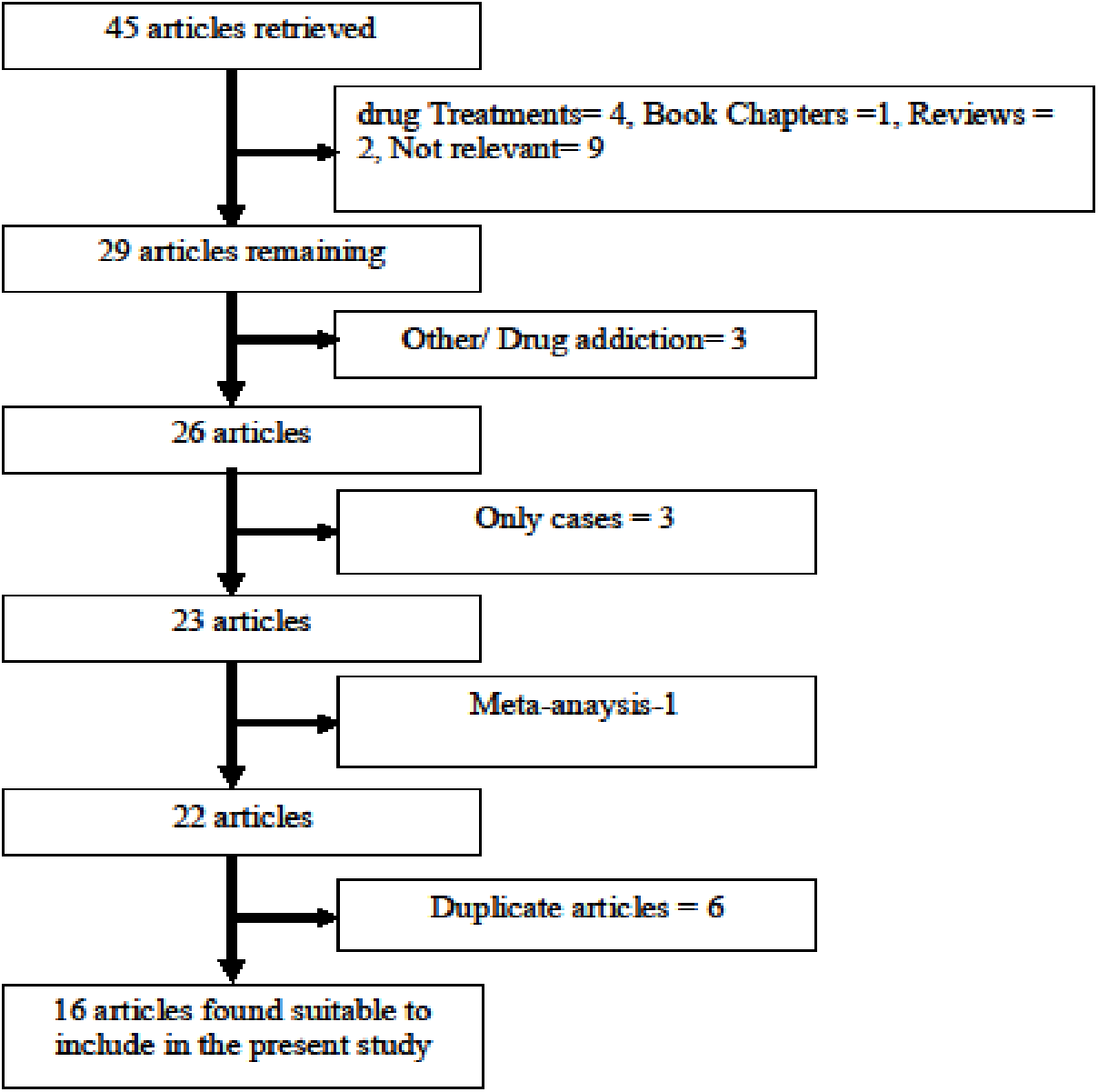
Flow Diagram of Study Search and Selection Process.

In all eighteen studies, total cases were 2278 with GG (766), AG (1056) and AA (456), and controls were 3717 with GG (1206), AG (1767), and AA (744) genotypes. In controls genotypes, percentage of GG, AG and AA were 32.45%, 47.54%, and 20.01% respectively (Figure 2). In total cases, genotype percentage of GG, AG, and AA was 33.62%, 46.36% and 20.02% respectively. In controls, the percentage of G and A alleles was 56.21 and 43.78 respectively (Figure 3). In eighteen studies included in the present meta-analysis, the highest sample size was 399 (Sery et al., 2006) and smallest case sample size was 28 (Carra et al., 2017).

**Figure 2.**
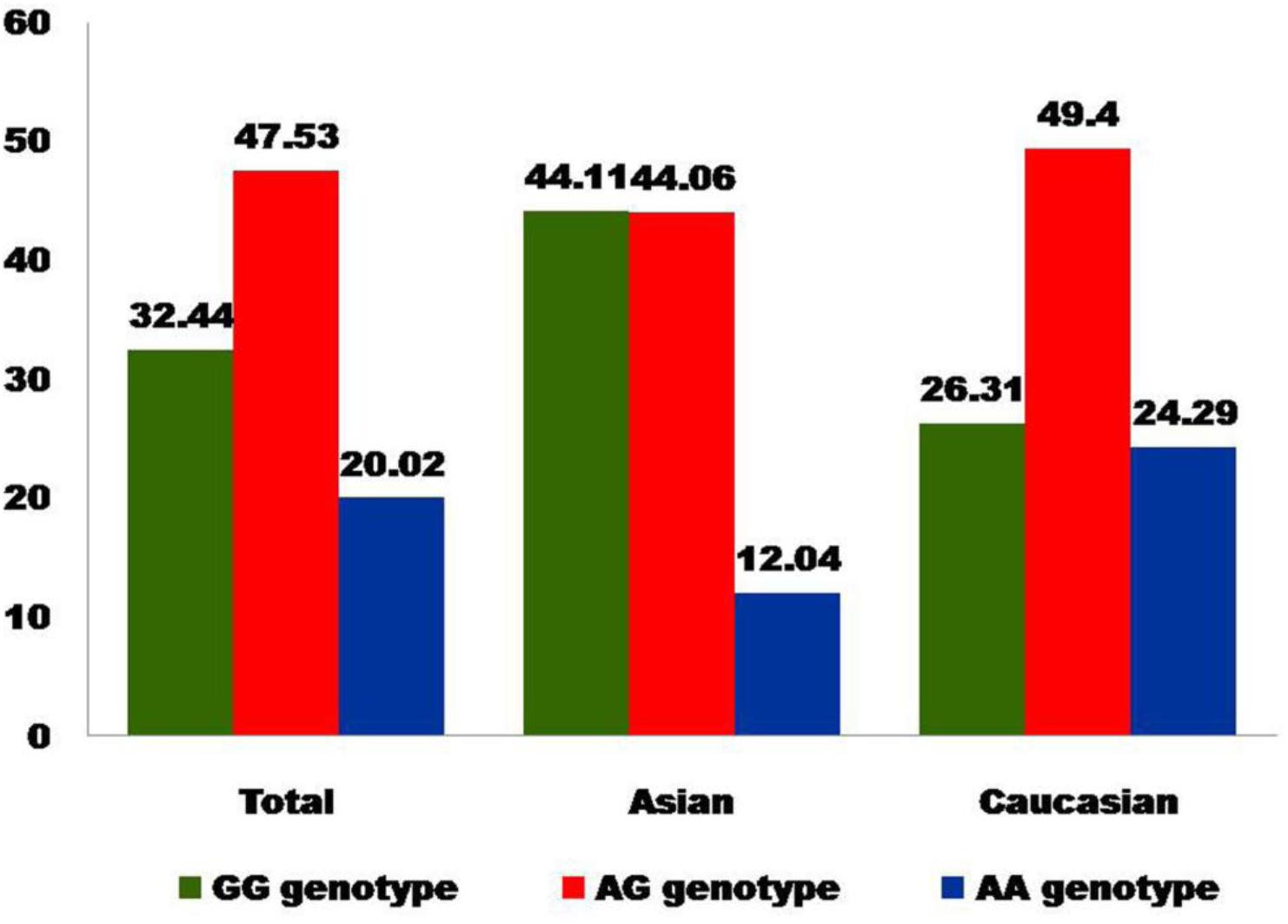
Bar diagram showing genotype percentage in control group of total studies, Asian studies and Caucasian studies

**Figure 3.**
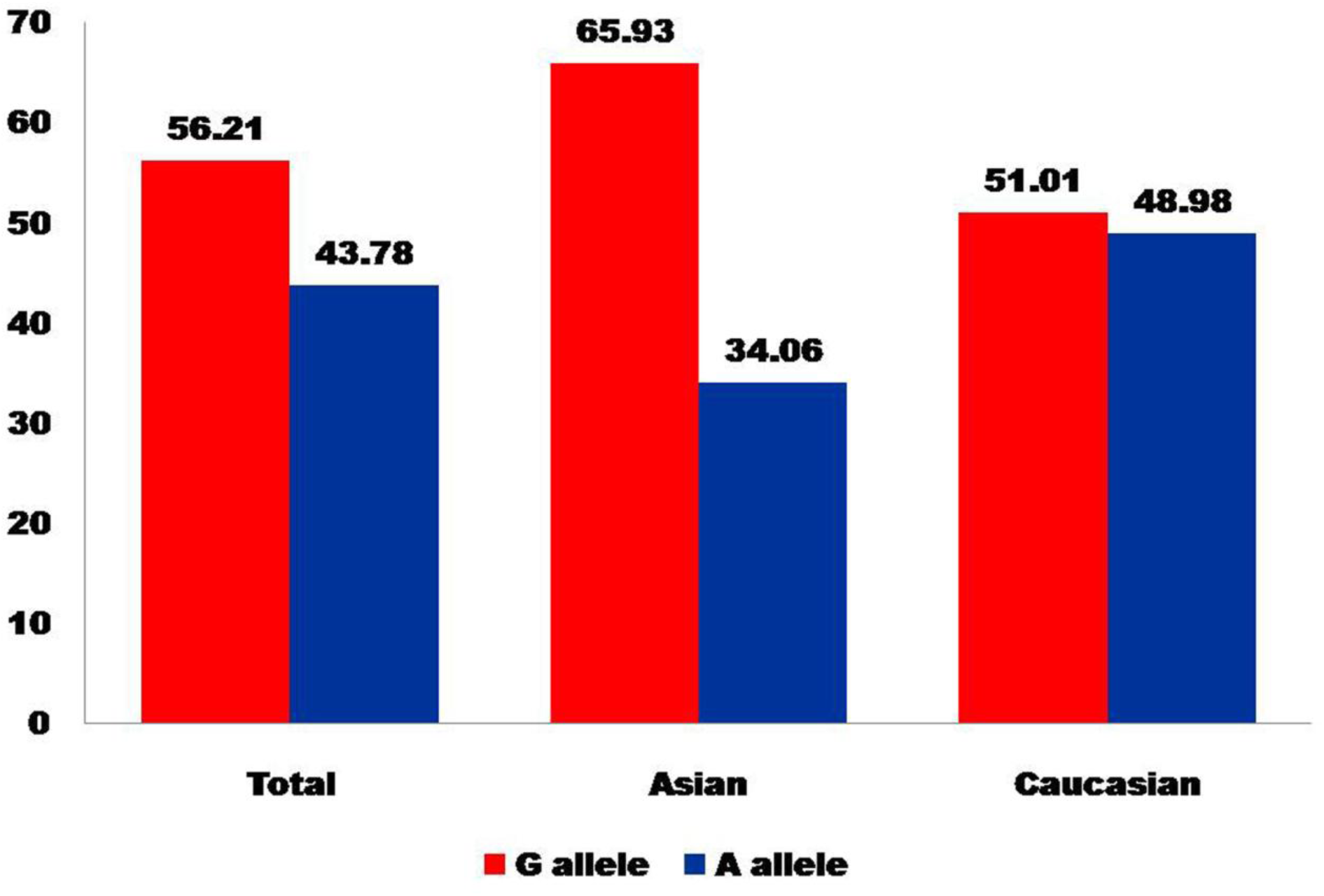
Bar diagram showing allele percentage in control group of total studies, Asian studies and Caucasian studies.

### Meta-analysis

All five genetic models; -allele contrast (A vs. G) homozygote (AA vs. GG), co-dominant (AG vs. GG), dominant (AA+AG vs. GG) and recessive (AA vs. AG+GG) models were used to evaluate Val158Met (G A) polymorphism as AD risk. Meta-analysis with allele contrast (A vs. G) did not show association with both fixed effect (A vs. G: OR= 1.007; 95%CI= 0.93-1.09; p= 0.84) and random effect model (A vs. G: OR= 1.02; 95%CI= 0.90-1.14; p= 0.003) (Table 2, Figure 4). There was observed no increased risk of AD using homozygote model (AA vs GG), with both fixed (AA vs GG: OR= 1.03; 95%CI= 0.87-1.21; p= 0.71) and random (AA vs GG: OR= 1.06; 95%CI= 0.81-1.38; p=0.69) effect models with high statistical heterogeneity between studies (Table 2). Association of mutant heterozygous genotype (AGvs.GG) was not observed significant with both fixed (AGvs.GG: OR= 0.98; 95%CI= 0.86-1.11; p= 0.80) and random (AGvs.GG:OR= 0.97; 95%CI= 0.86-1.11; p= 0.71) effect models. Combined mutant genotypes (AA+AG vs GG; dominant model) showed no positive association with AD using fixed (AA+AG vs GG: OR= 0.98; 95%CI= 0.87-1.10; p= 0.76)effect model (Table 2). Similarly the recessive genotypes model (AA vs. AG+GG) also did not show significant association with AD using both fixed (AA vs. AG+GG: OR= 1.4; 95%CI= 0.91-1.20; p= 0.49) and random (AA vs. AG+GG: OR= 1.04; 95%CI= 0.85-1.29; p= 0.61) effect models (Table 2).

**Table 2.**
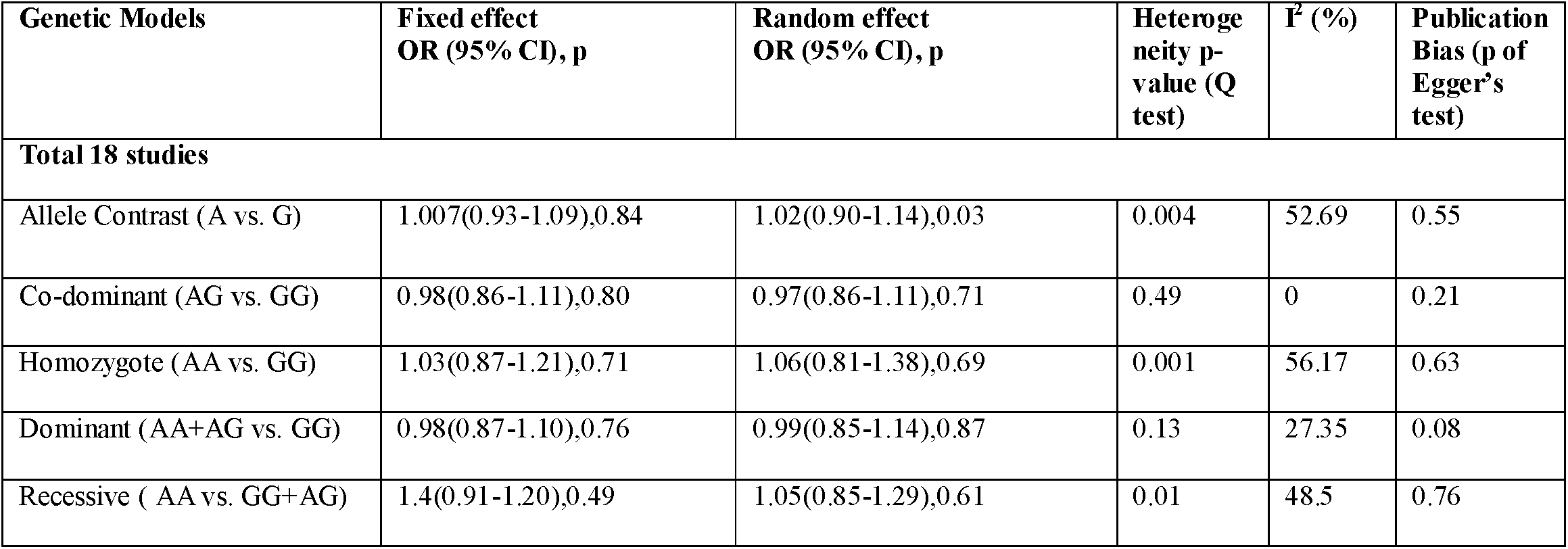
Summary estimates for the odds ratio (OR) in various allele/genotype contrasts, the significance level (p value) of heterogeneity test (Q test), and the I^2^ metric: overall analysis

| Genetic Models | Fixed effect<br>OR (95% CI), p | Random effect<br>OR (95% CI), p | Heterogeneity p-value (Q test) | I<sup>2</sup> (%) | Publication Bias (p of Egger's test) |
| --- | --- | --- | --- | --- | --- |
| **Total 18 studies** |  |  |  |  |  |
| Allele Contrast (A vs. G) | 1.007(0.93-1.09),0.84 | 1.02(0.90-1.14),0.03 | 0.004 | 52.69 | 0.55 |
| Co-dominant (AG vs. GG) | 0.98(0.86-1.11),0.80 | 0.97(0.86-1.11),0.71 | 0.49 | 0 | 0.21 |
| Homozygote (AA vs. GG) | 1.03(0.87-1.21),0.71 | 1.06(0.81-1.38),0.69 | 0.001 | 56.17 | 0.63 |
| Dominant (AA+AG vs. GG) | 0.98(0.87-1.10),0.76 | 0.99(0.85-1.14),0.87 | 0.13 | 27.35 | 0.08 |
| Recessive (AA vs. GG+AG) | 1.4(0.91-1.20),0.49 | 1.05(0.85-1.29),0.61 | 0.01 | 48.5 | 0.76 |

**Figure 4.**
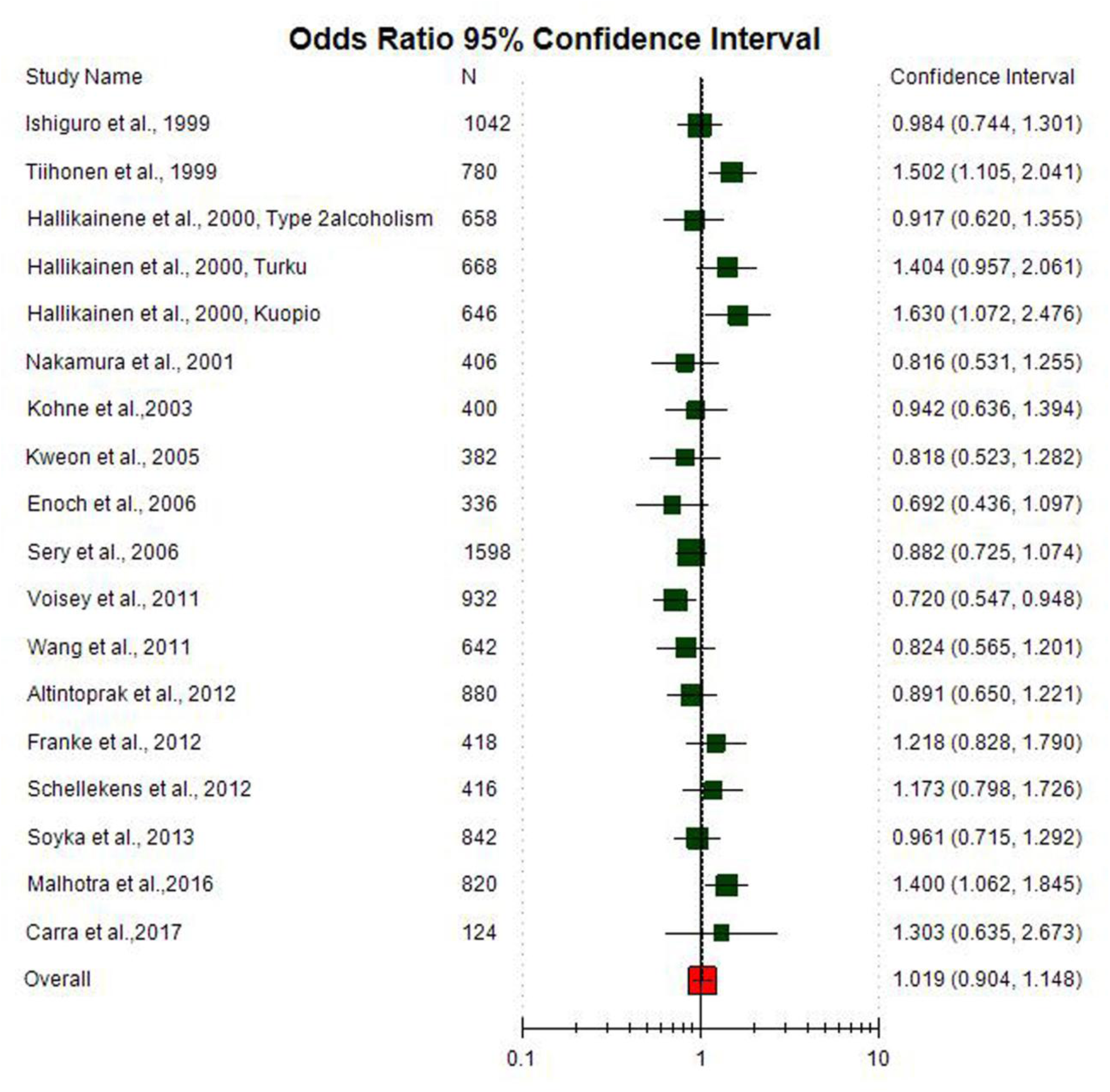
Random effect Forest plot of allele contrast model (AA vs. GG) of total 18 studies of COMT Val158Met(G472A polymorphism.

### Subgroup analysis and Heterogeneity

Subgroup analysis was done on the basis of ethnicity. The allele contrast meta-analysis of Asian studies did not show any significant association between COMT Val158Met polymorphism and AD (A vs. G: OR = 0.99; 95%CI= 0.86-1.13; p= 0.90; I^2^= 43.43%) (Figure 5) and meta-analysis of twelve Caucasian population based studies also did not show any association (A vs. G: OR = 1.02; 95%CI= 0.92-1.12; p= 0.75; I^2^=59.18%) (Figure 6).

**Figure 5.**
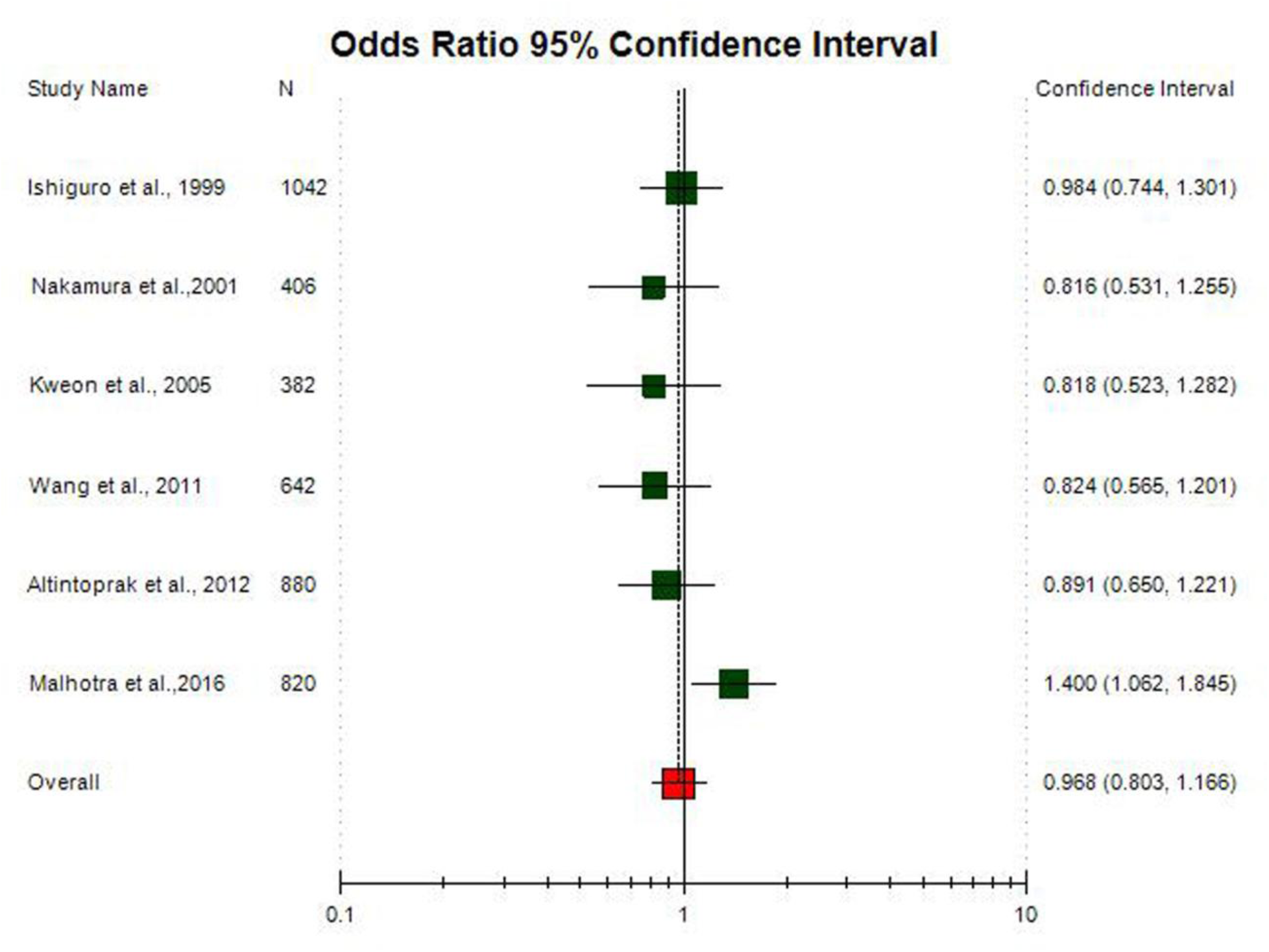
Random effect Forest plot of allele contrast model (A vs. G) of total 6 Asian studies of COMT Val158Met(G472A polymorphism.

**Figure 6.**
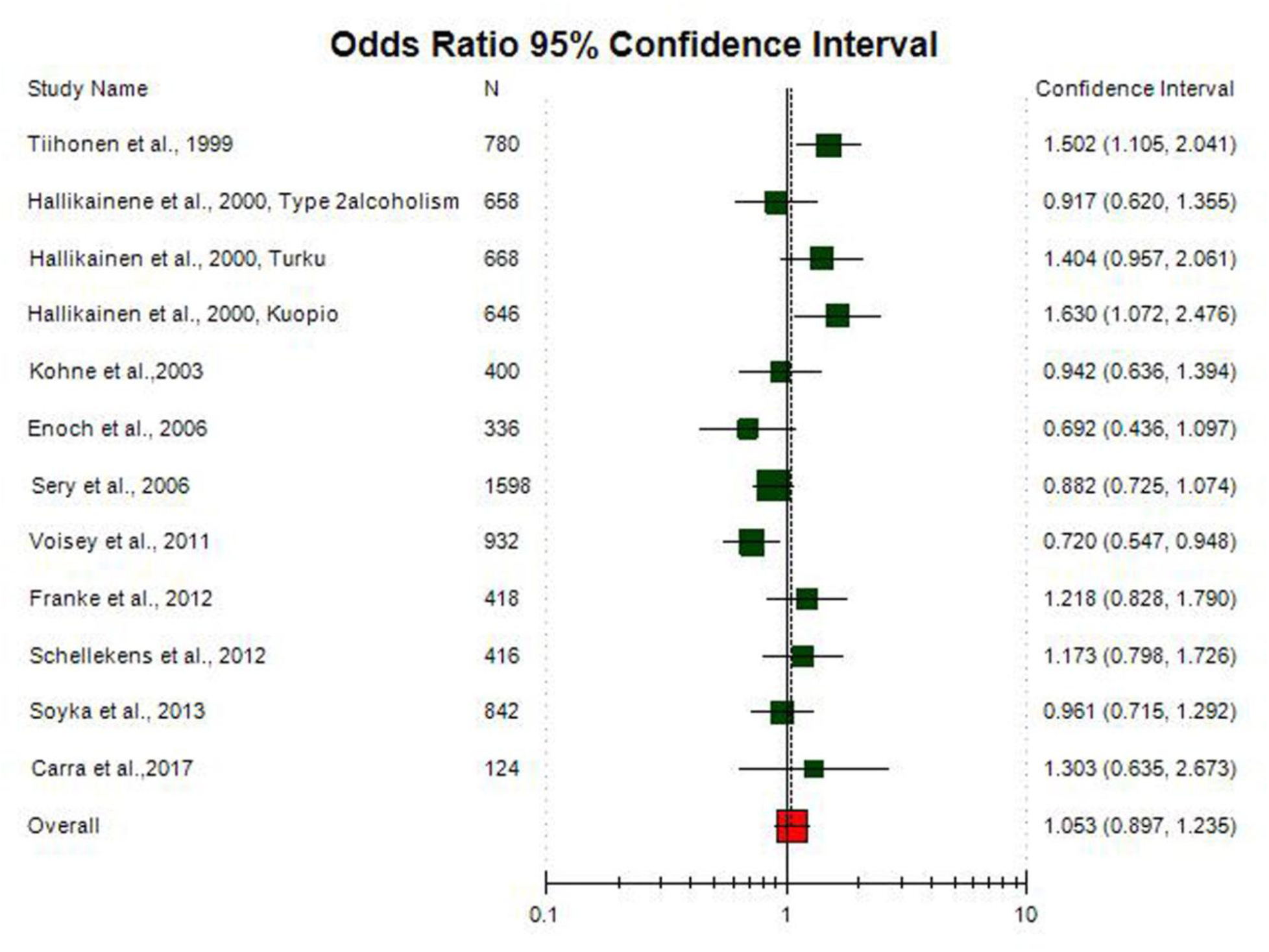
Random effect Forest plot of allele contrast model (A vs. G) of total 12 Caucasian studies of COMT Val158Met(G472A polymorphism.

A true heterogeneity existed between studies for allele contrast model (P_heterogeneity_=0.004, Q= 35.85, df= 17, I^2^= 52.69%, t^2^= 0.033) and homozygote model (P_heterogeneity_= 0.001, Q= 38.78, df= 17, I^2^= 56.17%, t^2^= 0.177) comparisons. The ‘I^2^’ value of more than 50% for between studies comparison in both allele contrast model and homozygote model shows high level of true heterogeneity. In Asian allele contrast meta-analysis showed low heterogeneity (P_heterogeneity_= 0.11, Q= 8.83, df= 5, I^2^= 43.43%, t^2^= 0.023), whereas Caucasian allele contrast meta-analysis showed significant high heterogeneity (P_heterogeneity_= 0.004, Q= 26.94, df= 11, I^2^= 59.187%, t^2^= 0.044).

### Publication bias

Publication bias was absent in all five genetic models and P value of Egger’s test was greater than 0.05 (A vs G, p= 0.55; AG vs GG, p= 0.21; AA vs GG, p= 0.63; AA+AG vs GG, p= 0.08; AA vs AG+GG, p= 0.76). Funnel plots using standard error and precision were also symmetrical (Figure 7).

**Figure 7.**
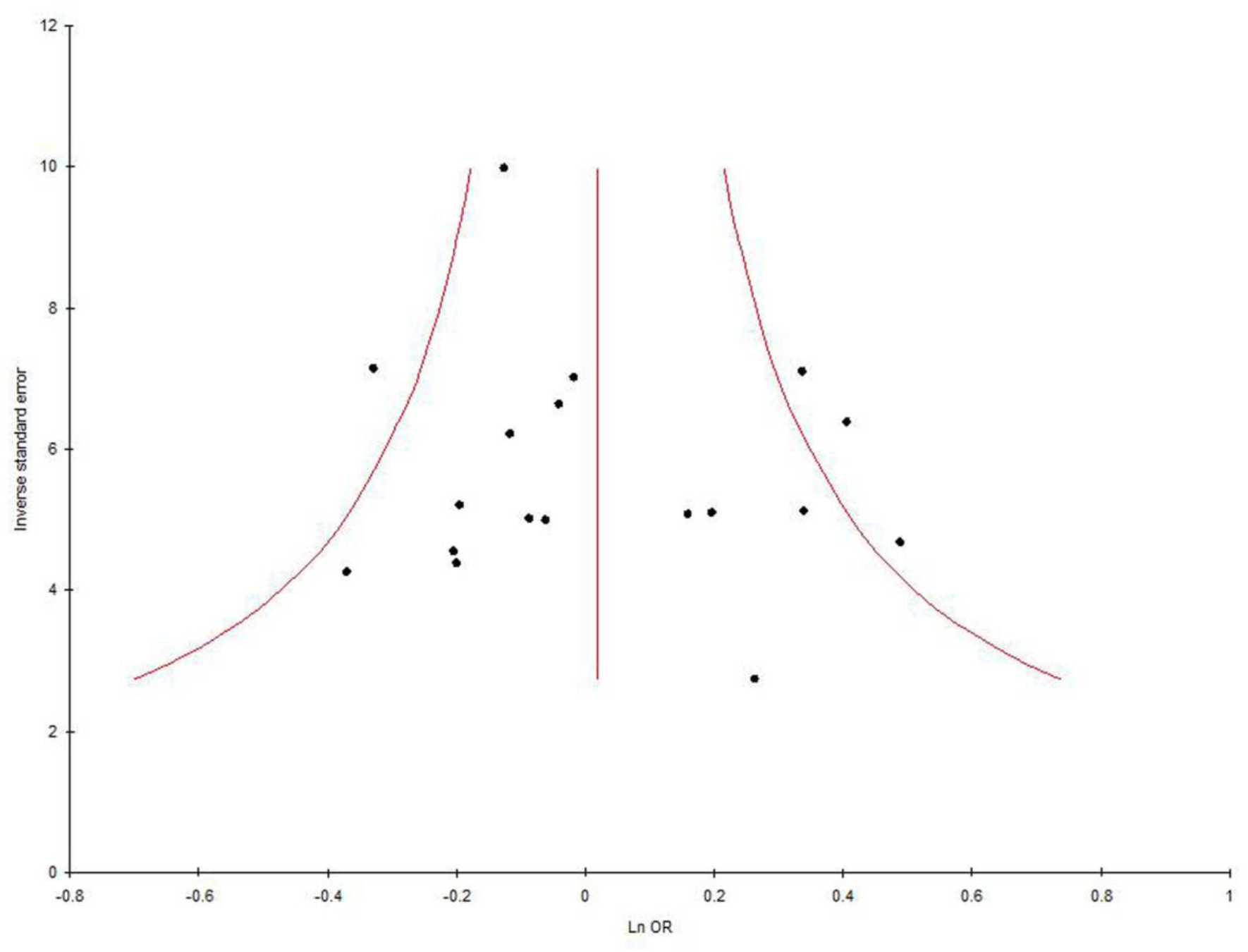
Funnel plot-Precision by log odds ratio for allele contrast model (A vs. G) of total 18 studies of COMT Val158Met(G472A polymorphism.

## Discussion

Current meta-analysis, which included 2278 AD cases and 3717 healthy controls, explored the associations between the maternal COMT Va158Met polymorphism and susceptibility to AD. Overall, we did not find any association between AA or AG genotypes with AD susceptibility. However, earlier COMT Val158Met (rs4680) genotype has been reported as risk factor for alcohol dependence, with a higher frequency of the Met allele in Type I alcohol-dependent patients (Tiihonen et al., 1999; Kweon et al. 2005). However, also negative association results have been reported (Hallikainen et al. 2000; Samochowiec et al. 2006).

Meta-analysis is a statistical tool, which overcome deficiencies of small studies /small sample analysis by combining data from several small studies and increasing the statistical power (lower type II error rate) (Ioannidis et., 2003). Several meta-analysis are published, which evaluated effects of polymorphism in susceptibility of diseases/disorders-down syndrome (Rai, 2011; Rai et al., 2017; Rai and Kumar, 2018), cleft lip and palate (Rai, 2014, 2017), Glucose-6-phosphate dehydrogenase deficiency (Kumar et al., 2016), male infertility (Rai and Kumar, 2017), schizophrenia (Yadav et al., 2015; Rai et al., 2017), obsessive compulsive disorder (Kumar and Rai, 2019), depression (Rai, 2014,2017), epilepsy (Rai and Kumar, 2018), Alzheimers disease (Rai, 2016,2017), esophageal cancer (Kumar and Rai, 2018), prostate cancer (Yadav et al., 2016), breast cancer (Rai et al., 2017), digestive tract cancer (Yadav et al., 2018), ovary cancer (Rai, 2016), endometrial cancer (Kumar et al., 2018), uterine leiomyioma (Kumar and Rai, 2018), MTHFR gene frequency (Yadav et al., 2018), and MTRR gene frequency (Yadav et al., 2019).

Present meta-analysis has several strength such as large number of studies (18 studies) along with larger sample size and absence of publication bias etc. Along with strength, meta-analysis has few imitations also, which should be acknowledged. like -(i) unadjusted OR was estimated, (ii) significant between-study heterogeneity was detected, (iii) single gene polymorphism (COMT Val158Met) was considered, (v) there might be gene-environment interaction for AD, however, we did not perform subgroup analysis due to lack of data on environmental factors. Pooled analysis of data from 18 separate studies from different ethnicities indicates that the COMT Val158Met polymorphism is not risk factor for AD. This association was complicated by higher between study heterogeneity. Future large-scale, different population-based association studies are required to investigate gene–environment interactions involving the COMT Val158Met polymorphism in determining AD risk.

## Data Availability

all data available in the manuscript.

## Notes

### Competing Interest Statement

The authors have declared no competing interest.

### Funding Statement

No funding

